# MALDI-TOF mass spectrometry of saliva samples as a prognostic tool for COVID-19

**DOI:** 10.1101/2021.12.10.21267596

**Authors:** Lucas C. Lazari, Rodrigo M. Zerbinati, Livia Rosa-Fernandes, Veronica Feijoli Santiago, Klaise F. Rosa, Claudia B. Angeli, Gabriela Schwab, Michelle Palmieri, Dmity J. S. Sarmento, Claudio R. F. Marinho, Janete Dias Almeida, Kelvin To, Simone Giannecchini, Carsten Wrenger, Ester C. Sabino, Herculano Martinho, José A. L. Lindoso, Edison L. Durigon, Paulo H. Braz-Silva, Giuseppe Palmisano

## Abstract

The SARS-CoV-2 infections are still imposing a great public health challenge despite the recent developments in vaccines and therapy. Searching for diagnostic and prognostic methods that are fast, low-cost and accurate is essential for disease control and patient recovery. The MALDI-TOF mass spectrometry technique is rapid, low cost and accurate when compared to other MS methods, thus its use is already reported in the literature for various applications, including microorganism identification, diagnosis and prognosis of diseases. Here we developed a prognostic method for COVID-19 using the proteomic profile of saliva samples submitted to MALDI-TOF and machine learning algorithms to train models for COVID-19 severity assessment. We achieved an accuracy of 88.5%, specificity of 85% and sensitivity of 91.5% for classification between mild/moderate and severe conditions. Then, we tested the model performance in an independent dataset, we achieved an accuracy, sensitivity and specificity of 67.18, 52.17 and 75.60% respectively. Saliva is already reported to have high inter-sample variation; however, our results demonstrates that this approach has the potential to be a prognostic method for COVID-19. Additionally, the technology used is already available in several clinics, facilitating the implementation of the method. Further investigation using a bigger dataset is necessary to consolidate the technique.

## Introduction

Since its emergence in Wuhan in December 2019, the virus responsible for the Coronavirus Disease-2019 (COVID-19), SARS-CoV-2, has spread worldwide and become a world-threatening disease^1^. According to the World Health Organization (WHO), by 17 of August of 2021 the COVID-19 pandemic reached a total of cumulative cases of 206,714,291 and cumulative deaths of 4,353,434 people world-wide^2^. Although many advances have been made in COVID-19 research, especially with the vaccines which initial data indicating its use was efficient and safe^3–5^, the emergence of SARS-CoV-2 variants with the increased transmission is concerning^6^. Thus, searching for new diagnosis and prognosis methods is still needed for increasing patient survival.

Regarding prognosis and diagnosis methods, simplicity, speed, low cost and accuracy are desirable qualities. Diagnosis methods for COVID-19 are present commonly in two forms, immunological assays or RT-qPCR; the former is a fast and cheap method, but can be very limited for early diagnosis since the immune response is still forming; the latter is considered the “gold standard” method and is the most used technique for COVID-19 diagnosis and disease tracking. However, the results are dependent of many factors, including proper sampling procedures and high-quality extraction kits ^7,8^. Variations on these methods have been implemented to achieve more accurate, affordable, easy to use and scalable diagnostic platforms ^9,10^. For being a relatively cheap and fast technique, MALDI-MS and machine learning algorithms have been implemented in many protocols, such as diagnosis and prognosis of several types of cancer ^11–13^, fungi and bacterial identification ^14,15^, detection of resistant fungi and bacteria ^16–18^, and COVID-19 diagnosis and prognosis ^19–23^. However, these works used mainly plasma, serum or nasal swab samples, while saliva samples are still poorly explored. The process for collecting saliva is simple, fast, and painless and require minimum supervision ^24^ saliva also contains high amounts of SARS-CoV-2 during the early stages of viral infection ^1^ and a recent study demonstrated that the salivary glands serve as a viral reservoir ^25^; thus, this biofluid can be useful as source material for diagnosis and prognosis of COVID-19. Taken together, the rapid turnaround time (typically 10 min), high accuracy (>95%) for detection of extremely low concentration of biomolecules, low cost in supplies and technical processing indicates MALDI-MS as an attractive option of choice in COVID-19 diagnosis and prognosis.

In this work, we propose a new method for COVID-19 diagnosis and prognosis using MALDI-TOF proteomic profile of salivary samples and machine learning. Samples from patients in a mild/moderate or severe conditions were used to train several machine learning algorithms and build a model to classify the samples into mild/moderate or severe COVID-19 cases. Moreover, another model was built to classify between infected and control samples. Thus, two models were trained one for diagnosis and one for prognosis, using a rapid and widespread technique, MALDI-TOF, which is common in clinical laboratories. Further investigations and a larger cohort are necessary to consolidate this technique as an alternative test for COVID-19 diagnosis or prognosis.

## Materials and Methods

### Materials

All reagents used were from Sigma Aldrich unless otherwise stated.

### Ethical statement

This study was conducted in accordance with the Declaration of Helsinki, and the protocol was approved by Research Ethics Committee of the “Instituto de Infectologia Emílio Ribas”, São Paulo, Brazil, protocol number CAAE 35589320.6.0000.0061. The invited volunteers were informed about the objectives, propositions and conditions of this project, in which those who agreed to participate in the research signed the free and informed consent term. Demographics, clinical data and samples were collected uniquely after the understanding of the study protocol and consent acknowledgement by the participants. A questionnaire on the health status of each participant was carried out. All participant information and samples were anonymized before use. Sample handling was carried out in a BSL2 laboratory.

### Individuals’ recruitment

COVID-19 infected-patients were classified in three groups according to the severity of the disease: 1) Mild form, characterized by the presence of flu-like symptoms, with the absence of dyspnoea and normal radiological examination. 2) Moderate, characterized by the presence of flu-like symptoms associated with pulmonary impairment < 50%, measured by computed tomography and O2 saturation >93% in room air. 3) Severe characterized by respiratory frequency greater than 30 breaths per minute, O2 saturation <93% in room air and pulmonary impairment >50% measured by computed tomography. During the period from January 13 to May 28, 2021 patients attending at the Instituto de Infectologia Emílio Ribas, Sao Paulo, Brazil, that tested molecularly positive for SARS-CoV-2 by nasopharyngeal swab were invited to enroll in the research study and provide saliva samples on the day of inclusion. Individuals under 18 years old and pregnant women were excluded.

Healthcare workers group (controls). Asymptomatic healthcare workers potentially exposed to patients or SARS-CoV-2 positive samples were invited to enrol into the study. Saliva collection was performed on the day of individual inclusion and tested negative for SARS-CoV-2 by RT-PCR.

### COVID-19 routine diagnostic method (RT-PCR)

A standard protocol was used for nasopharyngeal swab. Routine diagnosis protocol was applied for SARS-CoV-2 detection by RT-qPCR.

### Saliva sample preparation

Saliva samples were obtained from SARS-CoV-2 infected and healthy individuals by using a cotton pad device – Salivette™ (Sarstedt AG & CO. KG, Nümbrecht, Germany). The patients were asked to maintain the cotton in the mouth for 90 seconds; then, it was centrifuged at 1000 g for 5 minutes. After centrifugation, saliva samples were aliquoted and stored at −80 ºC. For viral inactivation, a solution of TFA 10% was added to the samples to obtain a final concentration of 1% of TFA in a BSL3 laboratory under all the safety measures needed. Then, protein quantification was performed using a Qubit assay (Thermo Fischer).

Different protein concentrations were evaluated before submitting the samples to MALDI-TOF analysis. Saliva samples were diluted with TFA 0.1% in the proportions of 1:1, 1:2, 1:5, 1:10 and 1:100; then, the samples were spotted directly in the MALDI target plate (Bruker Daltonics) using 1 μL of sample followed by 1 μL of matrix solution. Three different matrixes were also tested (sinapinic acid [SA], dihydroxybenzoic acid [DHB], and α-cyano-hydroxycinnamic acid [HCCA]) and the matrix solution was prepared by dissolving in acetonitrile/water 50:50 vol/vol containing 2.5 % TFA to obtain a concentration of 10 mg/mL.

Additionally, we tested if adding the same concentration of proteins for each sample in the MALDI plate would be better for group separation. For that, the protein quantification of all saliva samples was performed and a total of 0.2 μg of proteins (protein normalization) were added to the MALDI plate, this was compared with the direct spotting of the samples without taking into consideration the protein quantification.

### MALDI acquisition

Samples were analyzed in a MALDI-TOF Autoflex speed smartbeam mass spectrometer (Bruker Daltonics, Bremen, Germany) using FlexControl software (version 3.3, Bruker Daltonics). Spectra were recorded in the positive linear mode (laser frequency, 500 Hz; extraction delay time, 390 ns; ion source 1 voltage, 19.5 kV; ion source 2 voltage, 18.4 kV; lens voltage, 8.5 kV; mass range, 2400 to 20000 Da). Spectra were acquired using the automatic run mode to avoid subjective interference with the data acquisition. For each sample, 2500 shots, in 500-shot steps, were summed. All spectra were calibrated by using Protein Calibration Standard I (Insulin [M+H]+ = 5734.52, Cytochrome C [M+ 2H]2+ = 6181.05, Myoglobin [M+ 2H]2+= 8476.66, Ubiquitin I [M+H]+ = 8565.76, Cytochrome C [M+H]+ = 12 360.97, Myoglobin [M+H]+ = 16 952.31) (Bruker Daltonics, Bremen, Germany).

### Spectra processing

Data preprocessing and spectra evaluation were conducted using R packages. First, fid files were converted to mzML using MSconvert available in ProteoWizard (version: 3.0.20220) ^26^. The mzML files were loaded to R using MALDiquantForeign and processed using MALDIquant ^27^. The spectra for each sample were transformed (square root), smoothed (Savitzky-Golay and halfWindowSize of 10) ^28^ and the base line corrected using the TopHat algorithm ^29^. Then, the intensities were normalized using the total ion current and the peaks were selected with a signal-to-noise ratio of 2 and halfWindowSize of 10 ^22^. Peaks binning (tolerance of 0.003) and peak filtering (minimum frequency of 0.6) were performed for each group separately; then, a final binning (tolerance of 0.003) was performed with the groups merged. The resultant peaks were used to build an intensity matrix, which was further used for normality assessment (Shapiro-Wilk) and for a two-tailed Wilcoxon rank sum test corrected for multiple hypothesis testing using Benjamini-Hochberg. The peaks with a *P*-value < 0.05 were selected. The dataset was permuted 100 times and the global false discovery rate was calculated. For feature selection, the information gain method was used (FSelector package), since it is faster than wrapper methods and is classifier independent ^30^. Features with weight higher than 0 were selected.

### Machine learning

Six different algorithms were tested (SVM-P, GBM, SVM-R, NNET, NB, and RF) to classify samples in severe or mild/moderate and infected or control conditions. The training was carried through fourfold nested repeated ten times four-fold cross-validation using the Caret package, the data was split randomly. A random search was performed for hyperparameter tunning in the inner loop. The MLeval package was used to plot the ROC and PR curves. The AUC of ROC curves, accuracy, sensitivity and specificity were reported. Since the infected and control groups are not balanced, the balanced accuracy was used.

For the classification between severe and mild/moderate, the model with the best performance was selected for a validation step using another set of samples (test set) prepared and acquired separately from the training samples. The same preprocessing was carried out for the test set samples separately from the training samples. However, train and test sets were binned together to generate comparable features. SVM-R demonstrated the best performance in the algorithm comparison step; thus, it was selected for model training in the validation step. The parameters were fixed at sigma = 0.003329177 and C = 0.8915975, since they were the parameters that achieved the best accuracy for SVM-R. The model training was conducted in two ways; (1) The training set was used directly for model training, then, the test set was applied to the model and the metrics were calculated; (2) an extra pre-processing step was performed in the training set prior to model training, we conducted a feature selection using the information gain algorithm (FSelector) to select the features with higher importance (weights higher than zero). After feature selection, the model was trained using only the selected features, then the test set was applied to evaluate the model performance. A schematic workflow is presented in **Figure 1**.

**Figure 1:**
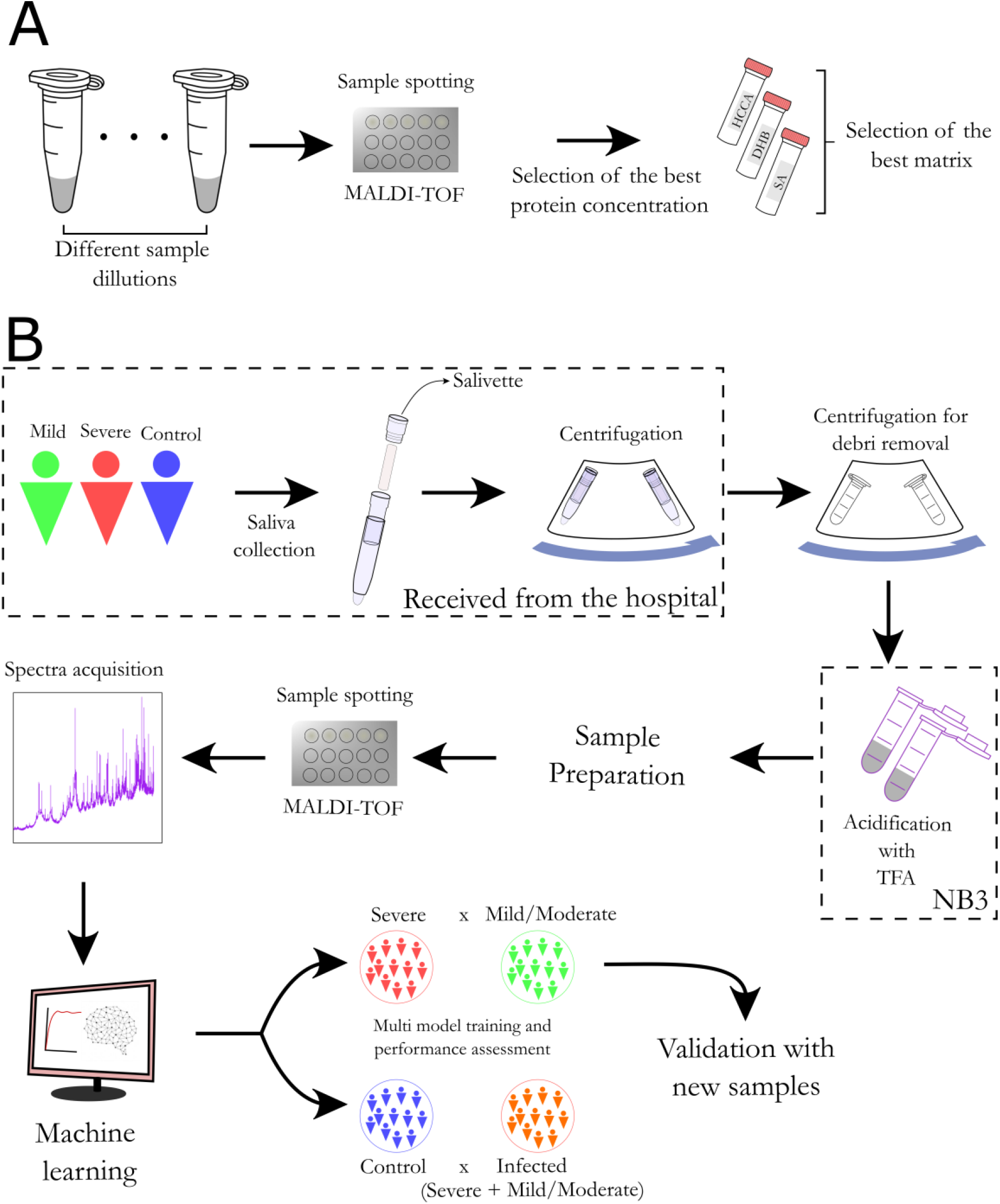
General workflow of the saliva sample preparation and MALDI-TOF analysis. (A) Method optimization. Initially, a range of different protein concentrations was evaluated to determine the optimal protein concentration. Subsequently, three MALDI matrices with better performances were evaluated and HCCA was selected as the optimal one. (B) The sample cohort consisted of four groups: severe (ICU), mild (emergency room), moderate (department) and control. The COVID-19 patients were confirmed by RT-qPCR of nasopharyngeal swab. Saliva samples were obtained using a salivette and virus inactivation was performed in an BSL-3 laboratory. Direct spotting of saliva on the MALDI plate was performed and the proteomic profile was acquired using a MALDI-TOF MS instrument. The MALDI-TOF spectra were pre-processed and analyzed in R environment, where statistically significant protein peaks were used to train multiple machine learning algorithms for group classification (training phase). The model with the best performances was used to analyze independent samples and calculate the accuracy, sensitivity and specificity of the model (validation phase).

## Results

In this study, we evaluated the potential of MALDI-TOF to identify prognostic biomarkers using saliva of COVID-19 patients. Initially, we tested a combination of analytical parameters to find the optimal conditions to obtain reliable and reproducible MS profiles (**Figure 1**). These conditions were optimized using a subset of samples belonging to the COVID-19 cohort. The entire cohort contained a total of 196 patients diagnosed with COVID-19, being 42 (21,4%) evaluated in emergency rooms, 81 (41,3 %) hospitalized and 73 (37,3%) in ICU and 36 controls. Within the COVID-19 patients, we classified the emergency room (outpatient and inpatient) patients as MILD/MODERATE condition and ICU patients as a SEVERE condition. Most patients were male with an average age of 52,4 years old, being 153 (78%) patients more than 40 years old. Most of them were not vaccinated (172, 87,7%) and used non-invasive oxygen support (106, 54%), 23 patients (11,7%) deceased due to COVID-19 complications. Most ICU patients were male and more than 40 years old, all patients with invasive oxygen support and 20 of the 23 death outcome patients were admitted to ICU. Sex (p=0.003), oxygen support type (p<0.001) and outcome (p<0.001) had statistical significance between emergency room/department and ICU. The most prevalent clinical symptoms were cough (67,9%), fever (61,7%), breathing difficulty (58,2%), fatigue (40,8%) and body or muscular aches (37,8%). We observed that sex (p=0.003), mechanical ventilation (p<0.001), body or muscular aches (p=0.046) and breathing difficulty (p=0.004) were statistically significant different between emergency room/department and ICU, being the latter the most prevalent in the patients treated in emergency rooms or hospitalized, and breathing difficulty most prevalent in ICU patients (**Table 1**). No statistical difference in sex and lower age for the controls was detected between COVID-19 patients and control subjects (**Supplementary Table 1**).

**Table 1:**
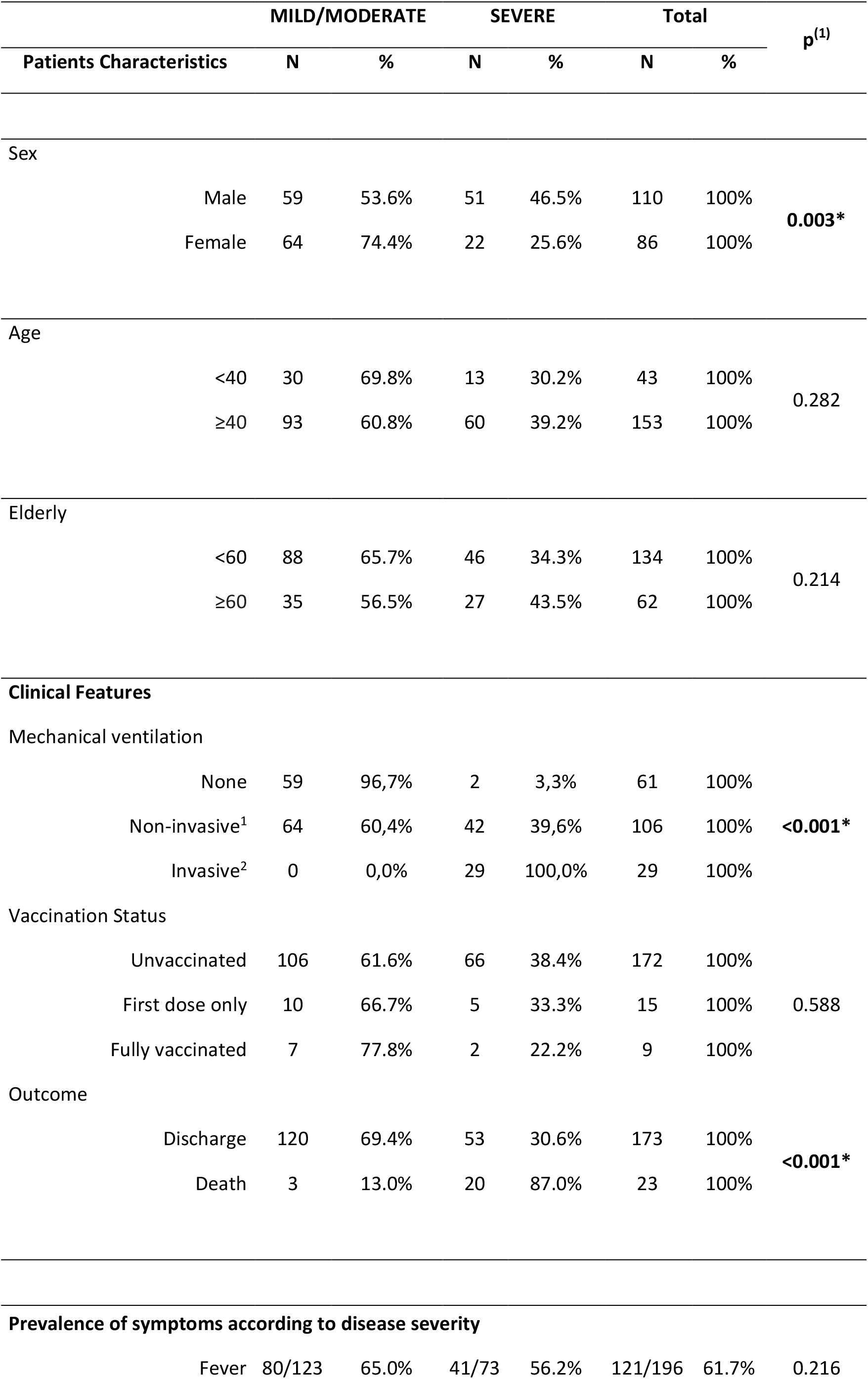
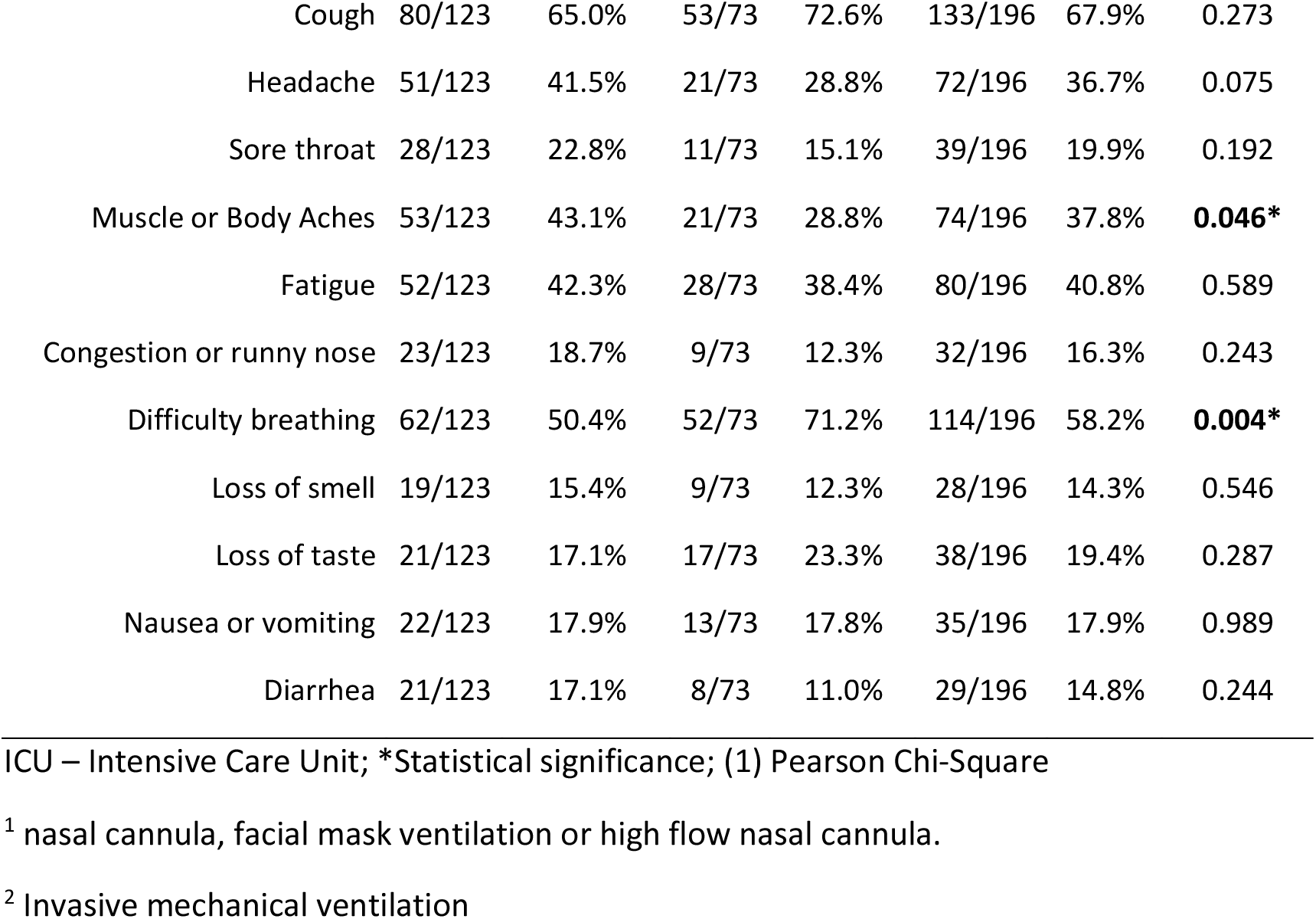
Clinical characteristics of the 192 COVID-19 patients investigated

|  |  | MILD/MODERATE |  | SEVERE |  | Total |  | p<sup>(1)</sup> |
| --- | --- | --- | --- | --- | --- | --- | --- | --- |
| Patients Characteristics |  | N | % | N | % | N | % |  |
| Sex |  |  |  |  |  |  |  |  |
|  | Male | 59 | 53.6% | 51 | 46.5% | 110 | 100% | 0.003* |
|  | Female | 64 | 74.4% | 22 | 25.6% | 86 | 100% |  |
| Age |  |  |  |  |  |  |  |  |
|  | <40 | 30 | 69.8% | 13 | 30.2% | 43 | 100% | 0.282 |
|  | ≥40 | 93 | 60.8% | 60 | 39.2% | 153 | 100% |  |
| Elderly |  |  |  |  |  |  |  |  |
|  | <60 | 88 | 65.7% | 46 | 34.3% | 134 | 100% | 0.214 |
|  | ≥60 | 35 | 56.5% | 27 | 43.5% | 62 | 100% |  |
| Clinical Features |  |  |  |  |  |  |  |  |
| Mechanical ventilation |  |  |  |  |  |  |  |  |
|  | None | 59 | 96,7% | 2 | 3,3% | 61 | 100% | <0.001* |
|  | Non-invasive<sup>1</sup> | 64 | 60,4% | 42 | 39,6% | 106 | 100% |  |
|  | Invasive<sup>2</sup> | 0 | 0,0% | 29 | 100,0% | 29 | 100% |  |
| Vaccination Status |  |  |  |  |  |  |  |  |
|  | Unvaccinated | 106 | 61.6% | 66 | 38.4% | 172 | 100% | 0.588 |
|  | First dose only | 10 | 66.7% | 5 | 33.3% | 15 | 100% |  |
|  | Fully vaccinated | 7 | 77.8% | 2 | 22.2% | 9 | 100% |  |
| Outcome |  |  |  |  |  |  |  |  |
|  | Discharge | 120 | 69.4% | 53 | 30.6% | 173 | 100% | <0.001* |
|  | Death | 3 | 13.0% | 20 | 87.0% | 23 | 100% |  |
| Prevalence of symptoms according to disease severity |  |  |  |  |  |  |  |  |
|  | Fever | 80/123 | 65.0% | 41/73 | 56.2% | 121/196 | 61.7% | 0.216 |

|  |  |  |  |  |  |  |  |
| --- | --- | --- | --- | --- | --- | --- | --- |
| Cough | 80/123 | 65.0% | 53/73 | 72.6% | 133/196 | 67.9% | 0.273 |
| Headache | 51/123 | 41.5% | 21/73 | 28.8% | 72/196 | 36.7% | 0.075 |
| Sore throat | 28/123 | 22.8% | 11/73 | 15.1% | 39/196 | 19.9% | 0.192 |
| Muscle or Body Aches | 53/123 | 43.1% | 21/73 | 28.8% | 74/196 | 37.8% | **0.046\*** |
| Fatigue | 52/123 | 42.3% | 28/73 | 38.4% | 80/196 | 40.8% | 0.589 |
| Congestion or runny nose | 23/123 | 18.7% | 9/73 | 12.3% | 32/196 | 16.3% | 0.243 |
| Difficulty breathing | 62/123 | 50.4% | 52/73 | 71.2% | 114/196 | 58.2% | **0.004\*** |
| Loss of smell | 19/123 | 15.4% | 9/73 | 12.3% | 28/196 | 14.3% | 0.546 |
| Loss of taste | 21/123 | 17.1% | 17/73 | 23.3% | 38/196 | 19.4% | 0.287 |
| Nausea or vomiting | 22/123 | 17.9% | 13/73 | 17.8% | 35/196 | 17.9% | 0.989 |
| Diarrhea | 21/123 | 17.1% | 8/73 | 11.0% | 29/196 | 14.8% | 0.244 |

ICU – Intensive Care Unit; \*Statistical significance; (1) Pearson Chi-Square
<sup>1</sup> nasal cannula, facial mask ventilation or high flow nasal cannula.
<sup>2</sup> Invasive mechanical ventilation

The optimization step consisted in testing several approaches for spectra acquisition. Initially, we tested the direct spotting method using several dilutions of the saliva. Three saliva samples were randomly selected from the cohort and analyzed in technical triplicates. We observed that direct spotting of samples diluted in a proportion of 1:1, 1:2, 1:5 and 1:10 presented similar number of peaks and maximum intensity, as well as a similar spectra profile (**Figure 2**). Thus, we selected the dilution of 1:10 that corresponded a protein concentration of approximately 0.2 μg/μL to conduct the following optimization step. The selection of optimal MALDI matrix was performed using the same saliva sample in technical triplicate. Three MALDI matrices, sinapinic acid [SA], dihydroxybenzoic acid [DHB], and α-cyano-hydroxycinnamic acid [HCCA] were tested and their MS profile compared. Different spectra profiles were obtained, being HCCA with higher maximum intensity, and DHB with more peaks identified (**Figure 2**). Although DHB had more peaks, we selected HCCA for the next analyses since it had lower variation in terms of number of peaks and maximum intensity. From these data it was possible to notice that analyses of technical replicates of the same saliva sample lead to less variation compared to different saliva samples (**Figure 2B–C vs 2E–F**).

**Figure 2:**
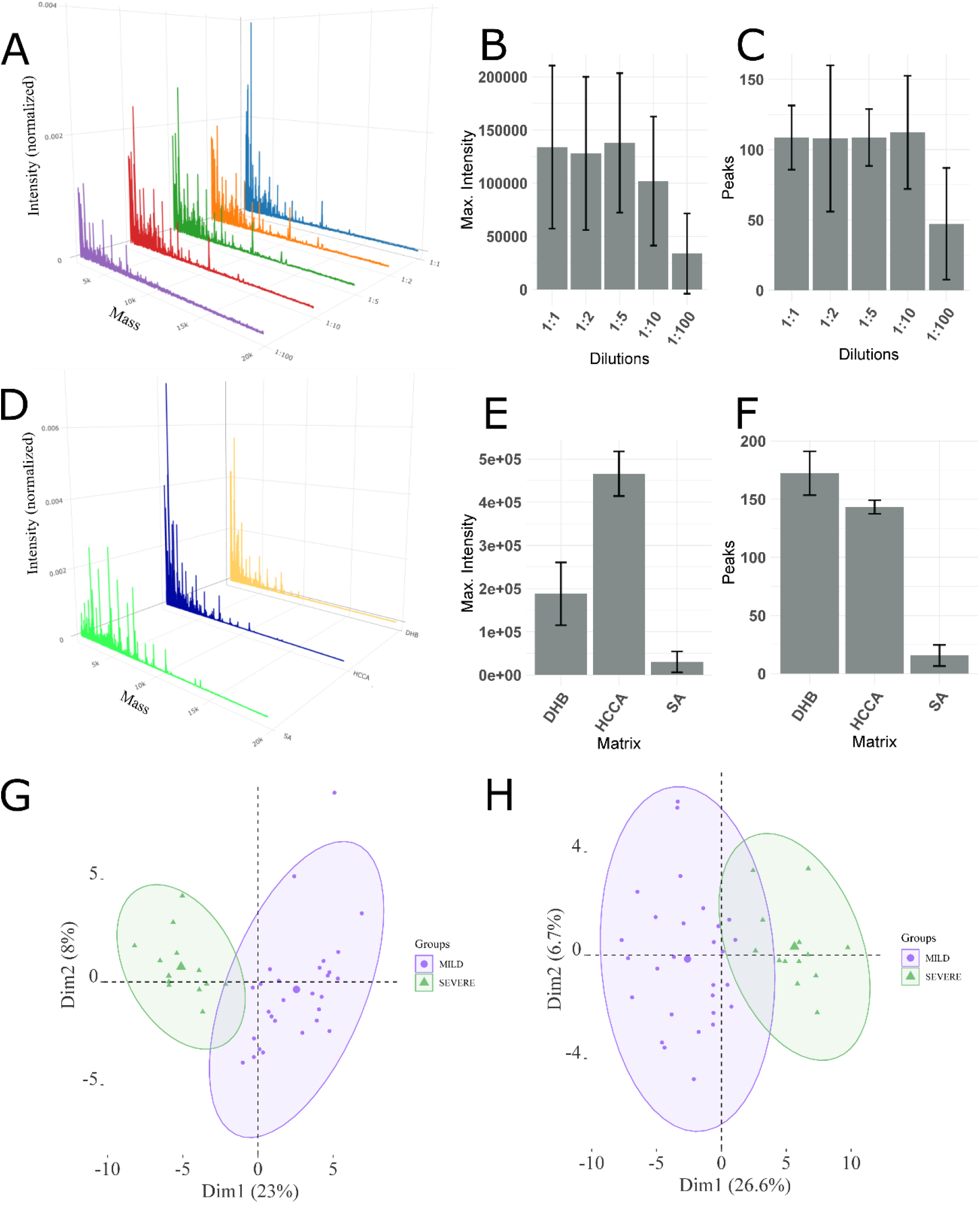
(A) Mean spectra profile obtained from three different samples in triplicate at different concentrations (1:1, 1:2, 1:5, 1:10 and 1:100 sample to TFA 0.1% ratio). (B) Maximum intensity of the spectra for each concentration. (C) The number of peaks identified for each concentration. (D) Mean spectra profile of one sample in triplicate for each matrix tested. (E) Maximum intensity of the spectra for each matrix. (F) Number of peaks detected for each matrix. (G) PCA of the samples that were analyzed with 0.2 μg of proteins. (H) PCA of the samples using 1ul of saliva irrespective of the protein concentration. In Figure 2G and 2H, MILD refers to MILD/MODERATE condition.

A small portion of the dataset (87 samples) was used to test whether loading the same amount of proteins in the MALDI plate would be better for group separation. We observed that loading 0.2 μg of proteins for each sample reduced the variation in the PCAs (**Figure 2G and H**). Thus, for the machine learning analysis we decided to conduct sample spotting of the entire dataset using 0.2 μg of proteins. The variation obtained between MILD/MODERATE and SEVERE was higher compared to the variation obtained within the same condition.

This study was divided into two parts comparing the saliva samples of: 1) control versus COVID-19 infected patients and 2) MILD/MODERATE versus SEVERE cases of COVID-19 patients. Average spectra for the Control (36 samples), MILD/MODERATE (51 samples), SEVERE (81 samples) and MILD/MODERATE+SEVERE (132 samples) are reported in **Figure 3A–D**.

**Figure 3:**
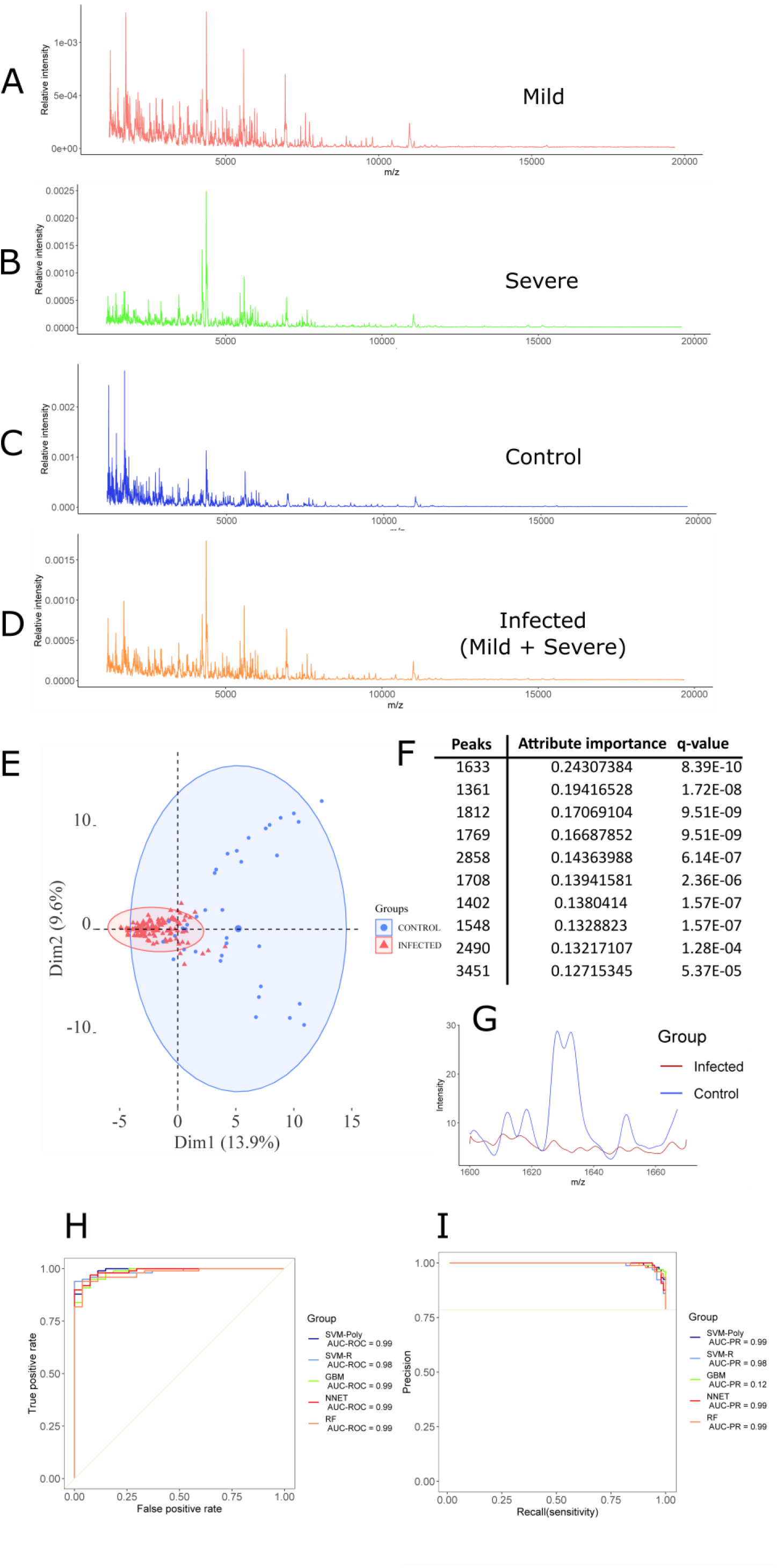
(A) MALDI-TOF MS mean spectrum of the saliva of the MILD/MODERATE group. (B) MALDI-TOF MS mean spectrum of the saliva of the SEVERE group. (C) MALDI-TOF MS mean spectrum of the saliva of the control group. (D) MALDI-TOF MS mean spectrum of the saliva of the infected group (MILD/MODERATE+SEVERE). (E) PCA for the control and infected groups. (F) The most ranked peaks after information gain filtering. (G) Spectra from the region of the highest ranked peak (1633). (H) Best ROC curves obtained from model training with control and infected samples. (I) Best PR curves obtained from model training with control and infected samples.

Initially, we aimed to comparing the MALDI-TOF MS profiles of control versus infected patients. A total of 132 samples of COVID-19 patients combining MILD, MODERATE and SEVERE were used as the infected group and 36 control samples were used as the control group. After peak filtration using the MALDIquant package, a total of 183 peaks were identified, which were reduced to 99 peaks after normality assessment and Wilcoxon rank sum test. The remaining peaks were used to plot the PCAs (**Figure 3E**) and train six machine learning models to classify between the conditions. The values for balanced accuracy and specificity were high for all models (**Supplementary Table 2**), with NB model having lower values. Sensitivity was high for SVM-P, SVM-R and RF, and balanced accuracy measures were similar for SVM-P and SVM-R, both with the highest value amongst the models tested, the complete list of all metrics obtained and all optimized hyperparameters for each fold is available at **Supplementary Table 3 and 4**. Overall, the best model was SVM-P, which scored a balanced accuracy, sensitivity and specificity of 92.9%, 92.2% and 93.6% The 99 statistically significant features (q value less than 0.05) were used to train the model (**Figure 3 H–I and Supplementary Table 5)**. Additionally, we performed an Information Gain filtering of the most relevant peaks and selected the 10 most ranked ones (**Figure 3 F**) and plotted the region of the most ranked peak (**Figure 3 G**).

These data showed a pronounced difference between the two conditions. This prompted us to investigate the MS profile of saliva samples from COVID-19 patients harboring different clinical characteristics.

Due to that, we analyzed 132 samples of COVID-19 patients (67% of the total samples) divided in 81 MILD/MODERATE and 51 SEVERE to identify MS-based saliva features. After peak filtration using MALDIquant package, a total of 139 peaks were identified, which were reduced to 44 peaks after normality assessment and Wilcoxon rank sum test. The remaining peaks were used to plot the PCA (**Figure 4A**) and train six machine learning models to classify between the conditions. The performance metrics for each model indicated that they differ, being the SVM-R the model with best mean accuracy (88.5%), mean sensitivity (91.5%) and mean specificity (85%) (**Figure 4F** and **Supplementary Table 6**). Also, specificity presented a high variation on every model tested, which was expected, since the saliva samples already presented a high variation in the PCA (**Figure 4A**). This was not observed in our previous study using the same platform but plasma as biofluid ^23^. The performance metrics in each fold for all six models and the optimized hyperparameters are available at the **Supplementary Table 7 and 8**. The best ROC and PR curves for each model are presented in the **Figure 4D–E**. The 44 statistically significant features (q value less than 0.05) were used to train the model **Figure 4D and Supplementary Table 9**. Additionally, we performed a feature selection by information gained to observe which were the most relevant peaks, the 10 most important peaks are reported in **Figure 4B** and the most rated peak is plotted in **Figure 4C**.

**Figure 4:**
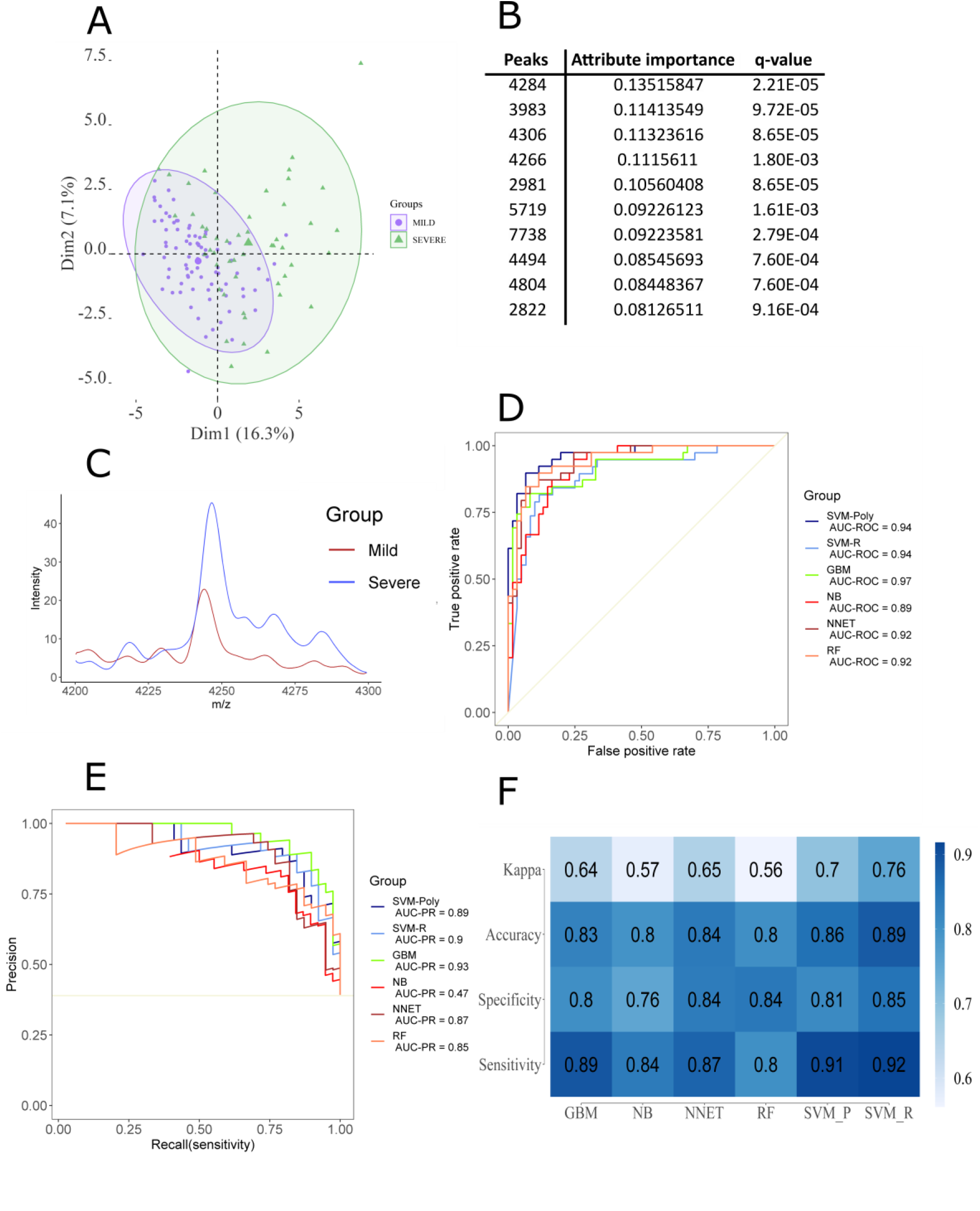
(A) PCA obtained through the analysis of the MILD/MODERATE and SEVERE conditions. (B) The 10 most relevant peaks were obtained through information gain filtering. (C) Comparison between MILD/MODERATE and SEVERE of the most discriminant peak. (D) Best ROC curves obtained from the model training. (E) Best PR curves obtained from the model training. (F) Mean accuracy, mean sensitivity, mean specificity and mean kappa were obtained in the model training step.

For SEVERE and MILD/MODERATE classification model validation, we prepared extra 64 samples (33% of the total samples) to test if the model could classify them correctly. We trained the SVR-R algorithm using optimized parameters (sigma = 0.003329177 and C = 0.8915975) retrieved from model selection step. First, we applied the test set to the trained model without feature selection, this approach yielded in an accuracy of 67.18%, a sensitivity of 52.17% and specificity of 75.60%. Then, we tested whether training the model using features that were selected by the information gain method could generate a better result; however, the metrics obtained were slightly lower, with an accuracy of 65.62%, sensitivity of 50% and specificity of 70.83%. The values obtained are expected since we used an independent dataset to validate the model generated during the training step. Moreover, being the MALDI-TOF MS features extracted from saliva samples highly variable, it is expected to retrieve lower accuracy, sensitivity and specificity values.

## Discussion

The use of machine learning to build models for COVID-19 diagnosis has been already reported in the literature. Several approaches have been tested, including the use of X-ray imaging data ^31^, emergency care admission exams ^32^ and routine blood tests ^33^. Also, mass spectrometry data have been used to train machine learning algorithms. Among the biofluids used and the mass spectrometry techniques performed we can list the analysis of nasopharyngeal swabs in MALDI-TOF spectrometer to build models for COVID-19 diagnosis ^22,34^, the analysis of proteomic profile of plasma samples by LC-MS/MS to build models for severity assessment and drug repurposing analysis ^35^, the use of the metabolomic profile obtained from plasma samples submitted to direct injection on high resolution MS system to build models for COVID-19 diagnosis and risk assessment ^21^, and the use of the proteomic profile of plasma samples submitted to MALDI-TOF to build models for COVID-19 risk assessment ^23^. Many of the studies are performed using biofluids that are invasive, such as plasma and nasopharyngeal swabs, and uses data acquisition techniques that are relatively expensive and time demanding. In order to slow down the disease spread, rapid and accurate identification of SARS-CoV-2 is necessary ^36^; thus, we describe here the use of a minimal invasive biofluid associated with MALDI-TOF, which is a relatively cheap and fast technique, for the diagnosis and prognosis of COVID-19 using machine learning algorithms. Our models attained a high performance, with the best model achieving an accuracy, sensitivity and specificity of 88.5%, 91.5% and 85% respectively for classifying between mild/moderate and severe cases of COVID-19; also, we achieved a balanced accuracy, sensitivity and specificity of 96.5%, 100% and 93.2% respectively for classifying between infected and control samples.

Although the models demonstrated a good performance in the training steps, the validation with independent data presented a lower performance than expected. It is known that the salivary proteome is a complex biofluid, with reports indicating an identification of 5500 proteins ^37^. Also, there are several factors that contribute to inter-individual variability of protein composition, such as taste stimulation, collection at different times of the day, age of infants, genetic polymorphism, and systemic pathologies ^38^. Saliva collection was conducted in a standardized manner; however, it is extremely difficult to control the behavior of each individual prior to sample collection. The direct spotting of the saliva samples might be another factor that contributed to the variability observed. Generally, direct spotting is more common in the literature, but other sample preparation methods might be applied prior spotting, such as protein digestion, glycoproteins enrichment, protein separation by electrophoretic gels, and peptide purification by C18 resins ^38–41^. Since our proposal was to develop the simplest yet accurate method for COVID-19 diagnosis, we did not evaluate the effect of more elaborate sample preparation methods; thus, further investigation is required in order to evaluate if the variability issue would be solved in the machine learning analysis.

We achieved a good accuracy for classifying between control and infected samples; however, it is important to note that our controls do not comprise samples obtained from patients that presented flu symptoms, meaning that it is still necessary to evaluate if the models trained are specific for COVID-19. Comparing infected patients with controls that presented similar symptoms is commonly done in the literature ^19,20,22^; however, since our study was aimed to prognosis, there were few control samples available in our cohort and all of them were from healthy patients. A study by Costa et al. (2021) demonstrated the potential of MALDI-TOF analysis of saliva samples and machine learning for COVID-19 diagnosis, they also reported high inter-individual variability and their models did not achieve sensitivity, specificity and accuracy values higher than 85% ^42^; our approach demonstrated a potential application for prognosis, and our models achieved similar performances, meaning that our results corroborates with what is expected for saliva samples.

This study demonstrates that MALDI-TOF MS and machine learning algorithms can be used to analyze saliva samples for prognosis purposes. This strategy is reproducible, easy to perform, fast, and low-cost. Since MALDI-TOF MS is already present in several clinical laboratories, this approach can be easily established in hospitals. However, it is important to note the limitations of this technique. We observed high inter-sample variation, which can reduce the performance of the models trained and reduce even more the performance during model generalization (validation with unseen data). This could be minimized by adding more samples to the cohort or by using more elaborate sample preparation methods. Therefore, a larger cohort should be analyzed to develop more robust models and inter-laboratory samples should be used to validate the findings. Moreover, improvements in the analytical part should be able to discriminate better between MILD and MODERATE groups.

## Data Availability

All data produced in the present study are available upon request to the authors.

## ACKNOWLEDGEMENTS

This work was supported by Fundação de Amparo à Pesquisa do Estado de São Paulo (FAPESP), GP (2018/18257-1, 2018/15549-1, 2020/04923-0), CW (2015/26722-8, 2017/03966-4), CRFM (2018/20468-0), PHB (2021/07490-0), ELD (2020/06409-1) and by Pró-Reitoria de Pesquisa da Universidade de São Paulo, PHB (2021.1.10424.1.9). GP (307854/2018-3), CW, and CRFM (302917/2019-5) were supported by Conselho Nacional de Desenvolvimento Científico e Tecnológico (CNPq). The funders had no role in study design. L Rosa-Fernandes, LC Lazari, RM Zerbinati and VF Santiago were supported by Coordenação de Aperfeiçoamento de Pessoal de Nível Superior (CAPES). M Palmieri was supported by Pró-Reitoria de Pesquisa da Universidade de São Paulo (2021.1.10424.1.9).

